# Nimodipine-Associated Standard Dose Reductions and Neurologic Outcomes After Aneurysmal Subarachnoid Hemorrhage: The Era of Pharmacogenomics

**DOI:** 10.1101/2024.05.16.24307178

**Authors:** Adriana Vázquez-Medina, Marion T. Turnbull, Courtney L. James, Jennifer B. Cowart, Elizabeth Lesser, Rickey E. Carter, Owen A. Ross, David A. Miller, James F. Meschia, Aixa De Jesús Espinosa, Richard Weinshilboum, W. David Freeman

## Abstract

Nimodipine, an L-type cerebroselective calcium channel antagonist, is the only drug approved by the US Food and Drug Administration for the neuroprotection of patients with aneurysmal subarachnoid hemorrhage (aSAH). Four randomized, placebo-controlled trials of nimodipine demonstrated clinical improvement over placebo; however, these occurred before precision medicine with pharmacogenomics was readily available. The standard enteral dose of nimodipine recommended after aSAH is 60 mg every 4 hours. However, up to 78% of patients with aSAH develop systemic arterial hypotension after taking the drug at the recommended dose, which could theoretically limit its neuroprotective role and worsen cerebral perfusion pressure and cerebral blood flow, particularly when concomitant vasospasm is present. We investigated the association between nimodipine dose changes and clinical outcomes in a consecutive series of 150 patients (mean age, 56 years; 70.7% women) with acute aSAH. We describe the pharmacogenomic relationship of nimodipine dose reduction with clinical outcomes. These results have major implications for future individualized dosing of nimodipine in the era of precision medicine.

## Introduction

Aneurysmal subarachnoid hemorrhage (aSAH) is a devastating condition caused by a ruptured intracranial aneurysm that carries a high 1-month mortality rate (10%-40%) and is commonly perceived as the worst headache of a person’s life. The worldwide annual incidence of aSAH is 1 per 10,000 patients per year, with an estimated 33,000 per year in the US. Approximately 55% of patients are younger than 55 when aSAH develops (1–3).

Following aSAH, 70% of patients develop cerebral arteriographic narrowing, or *vasospasm*, and 33% develop delayed cerebral ischemia (DCI) with secondary neurologic damage, with the latter leading to secondary cerebral infarction and additional neurologic disability (4). Clinically, vasospasm and DCI are both observed in a delayed fashion after aneurysm rupture, typically between 4 and 14 days after aSAH. The only neuroprotective drug approved by the US Food and Drug Administration (FDA) to treat patients with aSAH is nimodipine, a dihydropyridine, L-type cerebroselective calcium channel antagonist.

Although vasospasm and DCI are often confused for one another, they represent distinct cerebrovascular pathologies. For example, the FDA package insert indicates that nimodipine’s neuroprotective mechanism on outcomes is not correlated with preventing or relieving angiographic vasospasm (5). Therefore, nimodipine’s neuroprotective dose-response relationship with DCI and its effect on clinical outcomes remain poorly defined with regard to the drug’s pharmacometabolism. Further, a common adverse drug response (ADR) to nimodipine, systemic hypotension (systolic blood pressure <90 mm Hg), can necessitate reduction of the FDA-recommended dose of 60 mg (5–7).

All 4 randomized, placebo-controlled trials of nimodipine for patients with aSAH showed improvement in clinical outcomes compared to placebo (8–11). However, these randomized trials were conducted prior to the National Institutes of Health Human Genome Project in 2001 (12), which revolutionized the scientific approach to drug trials and led to increased use of precision medicine and pharmacogenomics in clinical practice. Dose reduction is common in clinical practice; therefore, understanding the pharmacogenomic implications of nimodipine--the only evidence-based medication for aSAH--is of paramount importance. In this study, we examine pharmacogenomics of cytochrome P450 (CYP) nimodipine drug metabolism in a consecutive series of patients with aSAH receiving the medication.

## Materials and Methods

After institutional review board (IRB 18-001392) approval, we retrospectively reviewed the clinical effects of hypotension and nimodipine dose reduction after aSAH admission and compared the available CYP genotypes in the electronic health record and an earlier prospective stroke genetics registry (13). The pharmacogenomics of nimodipine were reported in a small series of mainly healthy controls in Asia, where drug was shown to be metabolized predominantly by CYP3A4 and CYP3A5 enzymes (14). However, the study did not include patients with aSAH, the condition for which the drug is indicated (14). Therefore, we compared, when available, the CYP genotypes reported by Mayo Medical Laboratories, a Clinical Laboratory Improvement Amendments–approved genotyping facility to help clinicians assess the potential for drug-drug interactions.

Nimodipine is chiefly metabolized by the CYP enzyme system. The drug undergoes extensive hepatic metabolism by the isoforms CYP3A4, CYP3A5, and CYP2C19 subfamilies (15). We defined the genotype-phenotype pattern of nimodipine based on existing Mayo Medical Laboratories classification groupings (Figure 1), where allelic variants (particularly for CYP3A4 and CYP3A5 subtypes) convey the metabolic properties. The allelic combinations (Tables 1 and 2) categorize the metabolizers into the following groups: extensive, intermediate, average, and poor (11).

**Figure 1.**
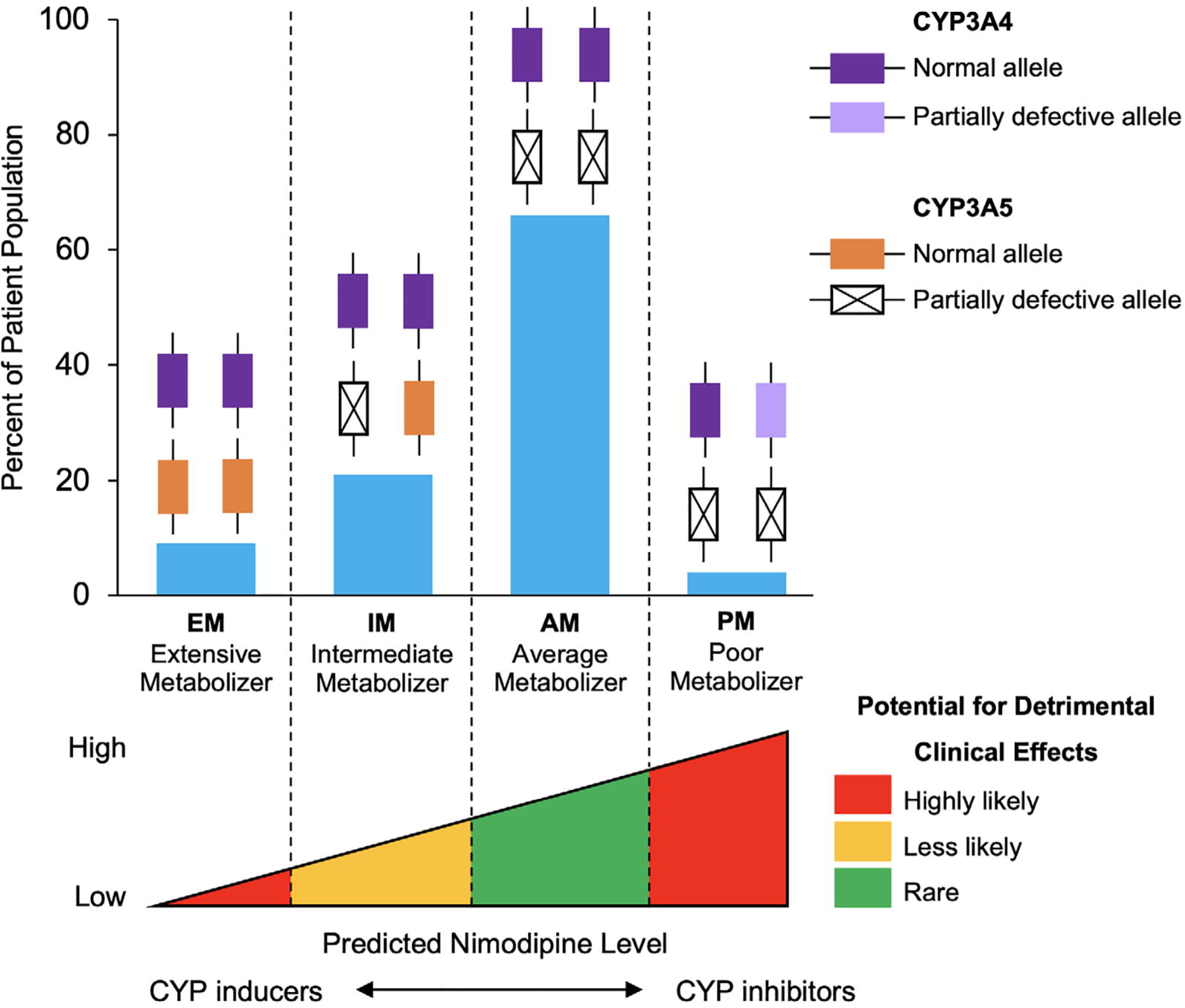
Proposed CYP3A4 and CYP3A5 Nimodipine Metabolizer Types as Extensive, Intermediate, Average, and Poor Metabolizers. CYP indicates cytochrome P450.

**Table 1.**
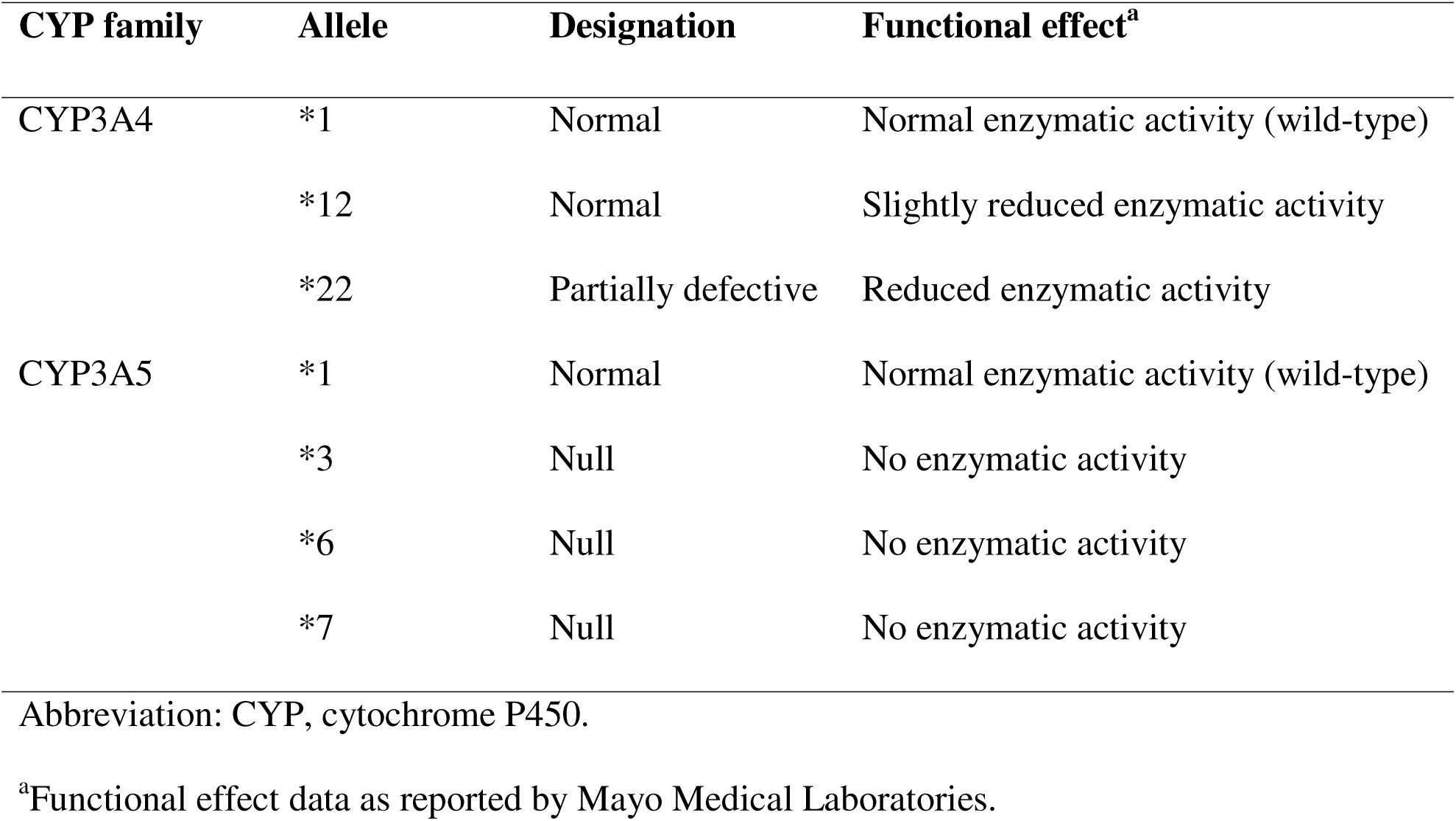
CYP Allelic Definitions and Functional Effects.

| CYP family | Allele | Designation | Functional effect<sup>a</sup> |
| --- | --- | --- | --- |
| CYP3A4 | *1 | Normal | Normal enzymatic activity (wild-type) |
|  | *12 | Normal | Slightly reduced enzymatic activity |
|  | *22 | Partially defective | Reduced enzymatic activity |
| CYP3A5 | *1 | Normal | Normal enzymatic activity (wild-type) |
|  | *3 | Null | No enzymatic activity |
|  | *6 | Null | No enzymatic activity |
|  | *7 | Null | No enzymatic activity |

Abbreviation: CYP, cytochrome P450.
<sup>a</sup>Functional effect data as reported by Mayo Medical Laboratories.

**Table 2.** CYP Genotype, Alleles, and Metabolizer Groupings and Predicted Effect on Nimodipine Serum.

| Metabolizer group | CYP3A4 | CYP3A5 | Predicted effect on nimodipine | Patient population, No. (%) |
| --- | --- | --- | --- | --- |
| Extensive | *1/*1 or *1/*12 | *1/*1 | Low levels | 13 (9) |
| Intermediate | *1/*1 | *3/*1, *1/*6, or *1/*7 | Reduced levels | 32 (21) |
| Average<sup>a</sup> | *1/*1 | *3/*3, *3/*7, *7/*7, or *3/*6 | Normal levels | 99 (66) |
| Poor | *1/*22 | *3/*3 or *3/*7 | High levels | 6 (4) |

Abbreviation: CYP, cytochrome P450.
<sup>a</sup>Average group is labeled as “normal levels” because it is the most common metabolizer group.

We defined clinical outcome as the patient’s modified Rankin scale (mRS) score at hospital discharge (16). We defined admission status using the World Federation of Neurologic Surgeons (WFNS) grade.

### Statistical Analysis

One-way ANOVA and Kruskal-Wallis rank sum tests were performed to compare numerical variables. For categorical variables, χ^2^ and Fisher tests were used accordingly. *P* values less than .05 were considered statistically significant, and all statistical tests were 2-sided. We generated a Sankey diagram using ChartExpo for Microsoft Excel (PPCexpo) to visualize dose changes during hospital stay and mRS score at discharge. Statistical analysis was performed using R Statistical Software, version 4.1.2 (R Foundation for Statistical Computing). Patients who did not have genotyping information in the electronic health record (n=2) were excluded.

## Results

To our knowledge, this is the largest pharmacogenomics investigation of CYP3A4 and CYP3A5 genotypes of nimodipine in patients with aSAH. Of the 152 patients with aSAH identified, 2 were excluded from analysis because their electronic health record did not contain pharmacogenomics registry information. Demographics and clinical characteristics for the 150 patients included are shown in Table 3. The mean age was 56.0, and most patients were women (70.7%), non-Hispanic or Latino (95.3%), White (66.7%), with history of hypertension (62%), and no history of diabetes (82.7%) or previous stroke (89.3%). Most aneurysms were in the anterior (26%) or posterior (21.3%) communicating artery. The most common surgical technique was aneurysmal coiling (76.7%). Table 2 shows the percentage of patients in each genotype-phenotype metabolizer group and allelic definitions for the enzymatic isoforms.

**Table 3.**
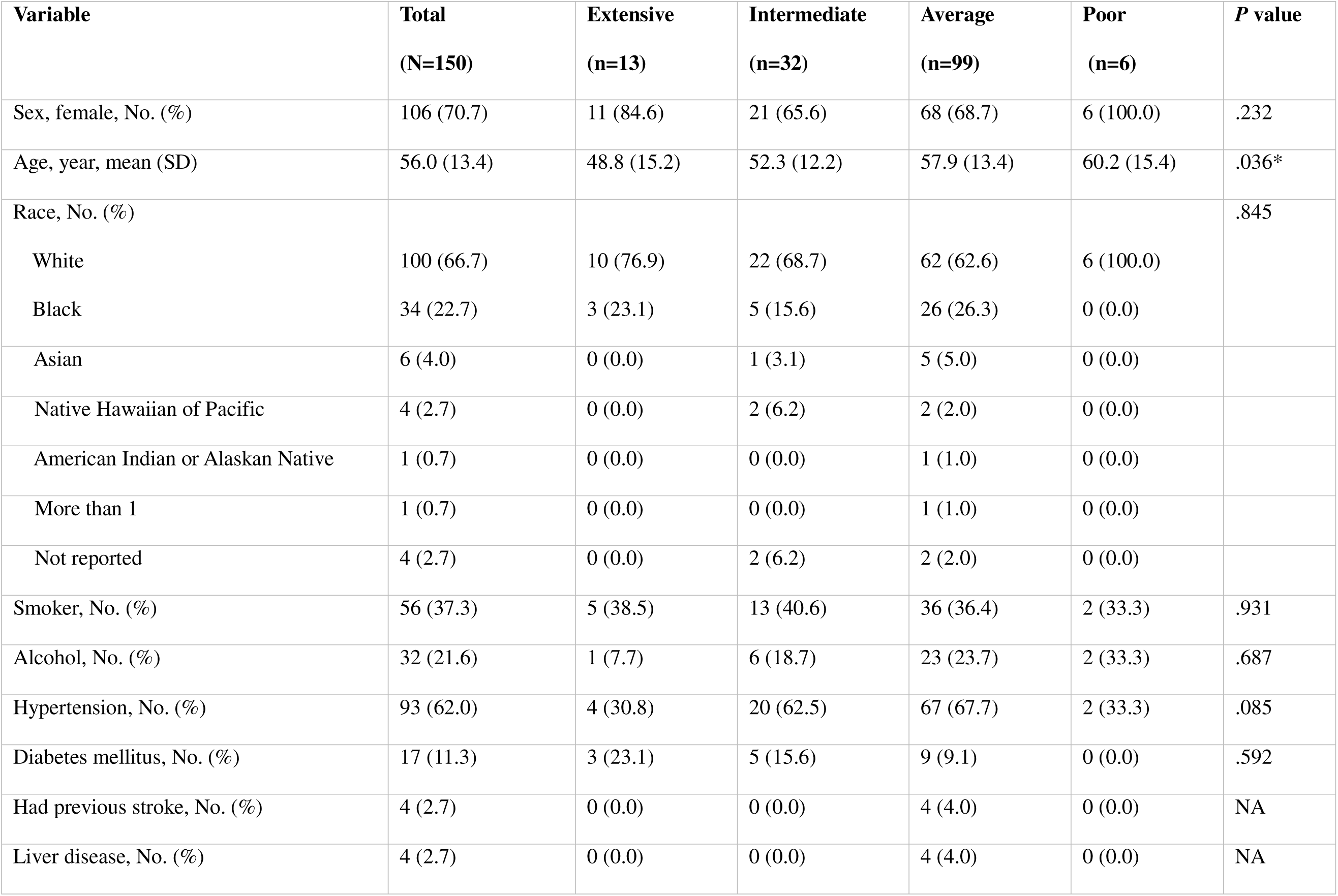
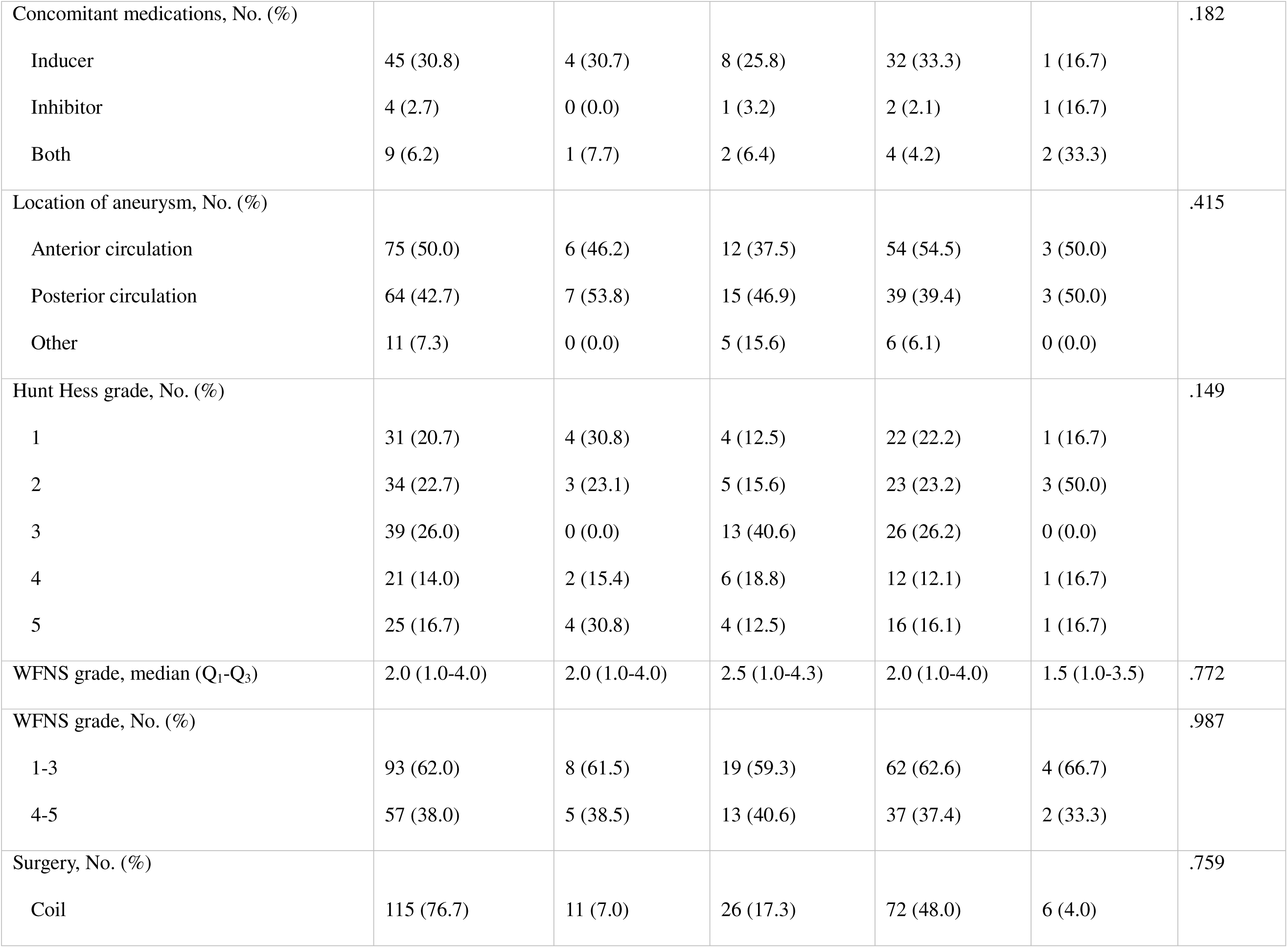
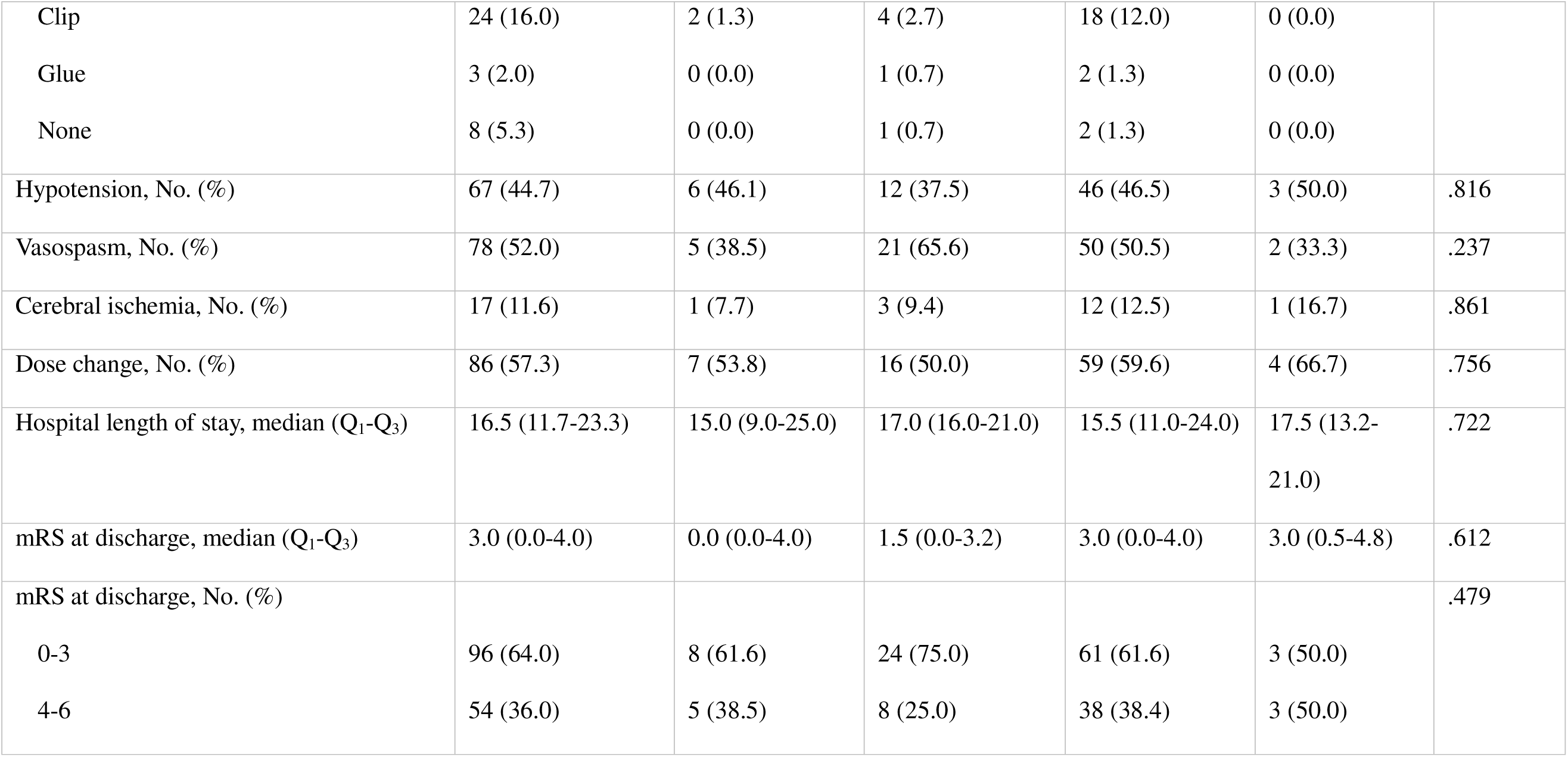
Comparison of Baseline Characteristics Based on Metabolizer Group.

| Variable | Total<br>(N=150) | Extensive<br>(n=13) | Intermediate<br>(n=32) | Average<br>(n=99) | Poor<br>(n=6) | P value |
| --- | --- | --- | --- | --- | --- | --- |
| Sex, female, No. (%) | 106 (70.7) | 11 (84.6) | 21 (65.6) | 68 (68.7) | 6 (100.0) | .232 |
| Age, year, mean (SD) | 56.0 (13.4) | 48.8 (15.2) | 52.3 (12.2) | 57.9 (13.4) | 60.2 (15.4) | .036* |
| Race, No. (%) |  |  |  |  |  | .845 |
| White | 100 (66.7) | 10 (76.9) | 22 (68.7) | 62 (62.6) | 6 (100.0) |  |
| Black | 34 (22.7) | 3 (23.1) | 5 (15.6) | 26 (26.3) | 0 (0.0) |  |
| Asian | 6 (4.0) | 0 (0.0) | 1 (3.1) | 5 (5.0) | 0 (0.0) |  |
| Native Hawaiian of Pacific | 4 (2.7) | 0 (0.0) | 2 (6.2) | 2 (2.0) | 0 (0.0) |  |
| American Indian or Alaskan Native | 1 (0.7) | 0 (0.0) | 0 (0.0) | 1 (1.0) | 0 (0.0) |  |
| More than 1 | 1 (0.7) | 0 (0.0) | 0 (0.0) | 1 (1.0) | 0 (0.0) |  |
| Not reported | 4 (2.7) | 0 (0.0) | 2 (6.2) | 2 (2.0) | 0 (0.0) |  |
| Smoker, No. (%) | 56 (37.3) | 5 (38.5) | 13 (40.6) | 36 (36.4) | 2 (33.3) | .931 |
| Alcohol, No. (%) | 32 (21.6) | 1 (7.7) | 6 (18.7) | 23 (23.7) | 2 (33.3) | .687 |
| Hypertension, No. (%) | 93 (62.0) | 4 (30.8) | 20 (62.5) | 67 (67.7) | 2 (33.3) | .085 |
| Diabetes mellitus, No. (%) | 17 (11.3) | 3 (23.1) | 5 (15.6) | 9 (9.1) | 0 (0.0) | .592 |
| Had previous stroke, No. (%) | 4 (2.7) | 0 (0.0) | 0 (0.0) | 4 (4.0) | 0 (0.0) | NA |
| Liver disease, No. (%) | 4 (2.7) | 0 (0.0) | 0 (0.0) | 4 (4.0) | 0 (0.0) | NA |
| Concomitant medications, No. (%) |  |  |  |  |  | .182 |
| Inducer | 45 (30.8) | 4 (30.7) | 8 (25.8) | 32 (33.3) | 1 (16.7) |  |
| Inhibitor | 4 (2.7) | 0 (0.0) | 1 (3.2) | 2 (2.1) | 1 (16.7) |  |
| Both | 9 (6.2) | 1 (7.7) | 2 (6.4) | 4 (4.2) | 2 (33.3) |  |
| Location of aneurysm, No. (%) |  |  |  |  |  | .415 |
| Anterior circulation | 75 (50.0) | 6 (46.2) | 12 (37.5) | 54 (54.5) | 3 (50.0) |  |
| Posterior circulation | 64 (42.7) | 7 (53.8) | 15 (46.9) | 39 (39.4) | 3 (50.0) |  |
| Other | 11 (7.3) | 0 (0.0) | 5 (15.6) | 6 (6.1) | 0 (0.0) |  |
| Hunt Hess grade, No. (%) |  |  |  |  |  | .149 |
| 1 | 31 (20.7) | 4 (30.8) | 4 (12.5) | 22 (22.2) | 1 (16.7) |  |
| 2 | 34 (22.7) | 3 (23.1) | 5 (15.6) | 23 (23.2) | 3 (50.0) |  |
| 3 | 39 (26.0) | 0 (0.0) | 13 (40.6) | 26 (26.2) | 0 (0.0) |  |
| 4 | 21 (14.0) | 2 (15.4) | 6 (18.8) | 12 (12.1) | 1 (16.7) |  |
| 5 | 25 (16.7) | 4 (30.8) | 4 (12.5) | 16 (16.1) | 1 (16.7) |  |
| WFNS grade, median (Q<sub>1</sub>-Q<sub>3</sub>) | 2.0 (1.0-4.0) | 2.0 (1.0-4.0) | 2.5 (1.0-4.3) | 2.0 (1.0-4.0) | 1.5 (1.0-3.5) | .772 |
| WFNS grade, No. (%) |  |  |  |  |  | .987 |
| 1-3 | 93 (62.0) | 8 (61.5) | 19 (59.3) | 62 (62.6) | 4 (66.7) |  |
| 4-5 | 57 (38.0) | 5 (38.5) | 13 (40.6) | 37 (37.4) | 2 (33.3) |  |
| Surgery, No. (%) |  |  |  |  |  | .759 |
| Coil | 115 (76.7) | 11 (7.0) | 26 (17.3) | 72 (48.0) | 6 (4.0) |  |
| Clip | 24 (16.0) | 2 (1.3) | 4 (2.7) | 18 (12.0) | 0 (0.0) |  |
| Glue | 3 (2.0) | 0 (0.0) | 1 (0.7) | 2 (1.3) | 0 (0.0) |  |
| None | 8 (5.3) | 0 (0.0) | 1 (0.7) | 2 (1.3) | 0 (0.0) |  |
| Hypotension, No. (%) | 67 (44.7) | 6 (46.1) | 12 (37.5) | 46 (46.5) | 3 (50.0) | .816 |
| Vasospasm, No. (%) | 78 (52.0) | 5 (38.5) | 21 (65.6) | 50 (50.5) | 2 (33.3) | .237 |
| Cerebral ischemia, No. (%) | 17 (11.6) | 1 (7.7) | 3 (9.4) | 12 (12.5) | 1 (16.7) | .861 |
| Dose change, No. (%) | 86 (57.3) | 7 (53.8) | 16 (50.0) | 59 (59.6) | 4 (66.7) | .756 |
| Hospital length of stay, median (Q<sub>1</sub>-Q<sub>3</sub>) | 16.5 (11.7-23.3) | 15.0 (9.0-25.0) | 17.0 (16.0-21.0) | 15.5 (11.0-24.0) | 17.5 (13.2-21.0) | .722 |
| mRS at discharge, median (Q<sub>1</sub>-Q<sub>3</sub>) | 3.0 (0.0-4.0) | 0.0 (0.0-4.0) | 1.5 (0.0-3.2) | 3.0 (0.0-4.0) | 3.0 (0.5-4.8) | .612 |
| mRS at discharge, No. (%) |  |  |  |  |  | .479 |
| 0-3 | 96 (64.0) | 8 (61.6) | 24 (75.0) | 61 (61.6) | 3 (50.0) |  |
| 4-6 | 54 (36.0) | 5 (38.5) | 8 (25.0) | 38 (38.4) | 3 (50.0) |  |

A simplification of the heterogeneity of multiple dose changes (ie, 60 mg, 30 mg, 15 mg, or 0 mg) in the cohort from hospitalization to discharge is reported in Figure 2. All patients initially received the FDA-recommended nimodipine dose of 60 mg every 4 hours, and 42.7% of patients did not have their dose reduced from the standard. When reduced, doses were either left at the reduced level (30 mg [25.3%], 15 mg [8.0%], or 0 mg [0.67%]) or returned to 60 mg prior to discharge (23.3%). The most common mRS score at discharge was 0 (Figure 2). The distribution of mRS scores at discharge for patients with reduced dose or no dose change is displayed in Figure 3. Figure 4 shows mRS scores by metabolizer group.

**Figure 2.**
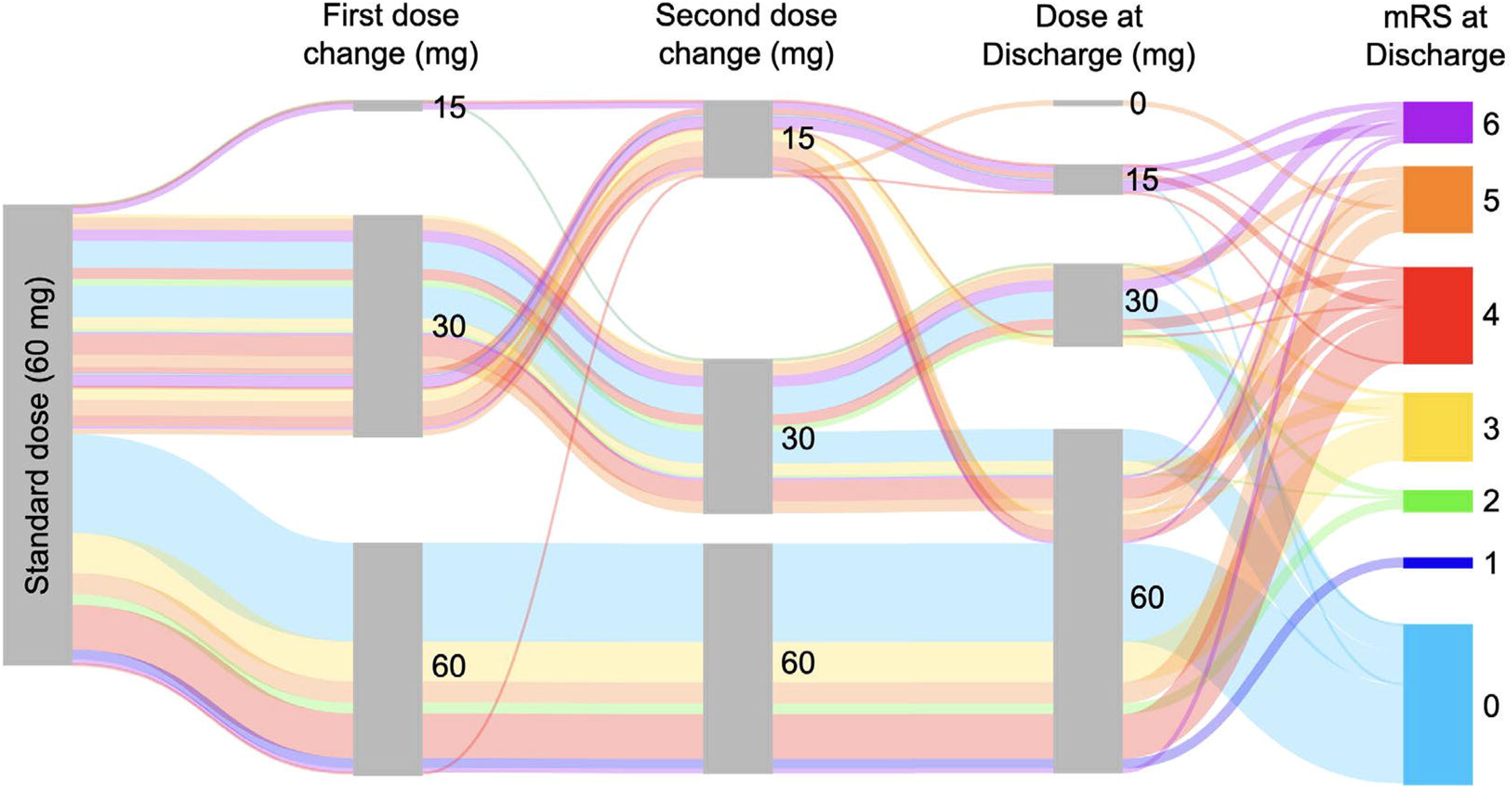
Sankey Flow Diagram Showing Heterogeneity of Multiple Dose Changes (60 mg, 30 mg, 15 mg, or 0 mg) in aSAH Patients During Hospital Stay and Until Discharge. At discharge, an mRS score is assigned as a predictor of outcome. hrs indicates hours; mRS, modified Rankin scale.

**Figure 3.**
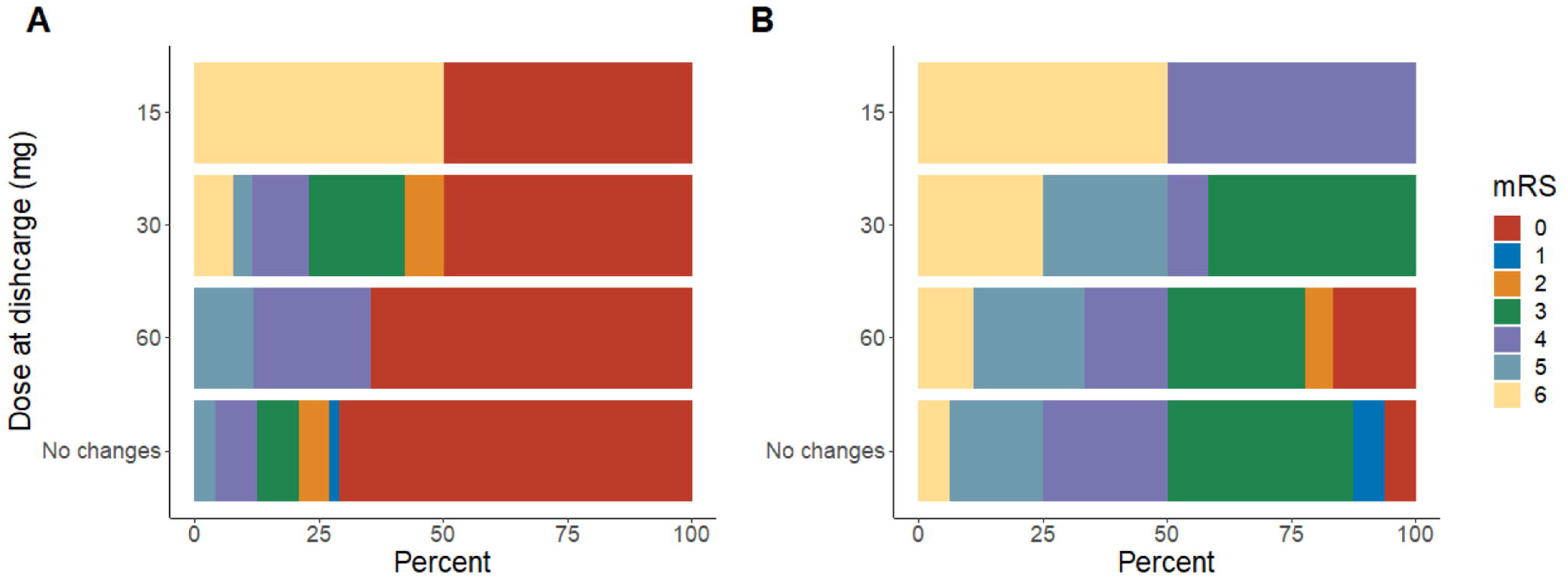
Distribution of mRS Scores at Discharge Among Dose Reduced Patients. A, mRS scores for patients with reduced dose or no dose change at discharge who had WFNS grades of 1-3. B, mRS scores for patients with reduced dose or no dose change at discharge who had WFNS grades of 4-5. mRS indicates modified Rankin scale; WFNS, World Federation of Neurological Surgeons.

**Figure 4.**
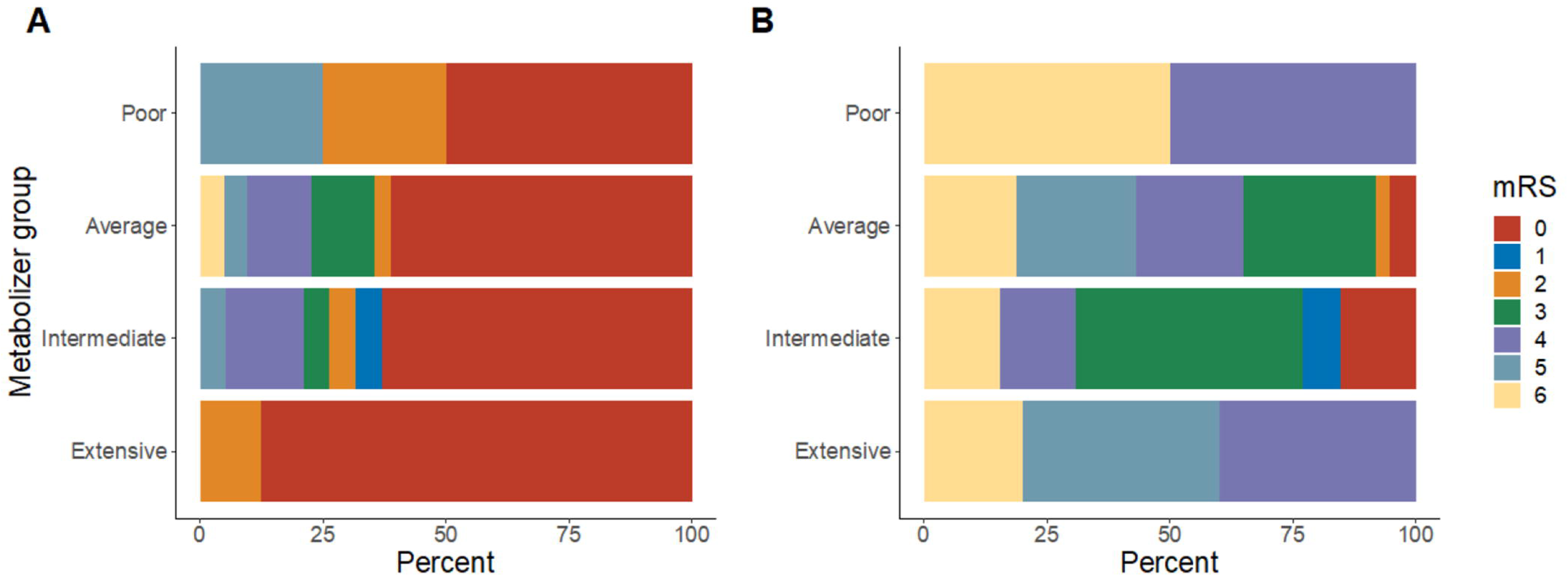
Distribution of mRS Scores at Discharge Among the Different Metabolizer Groupings. A, mRS outcome for each metabolizer grouping at discharge among patients with WFNS grades of 1-3. B, mRS outcome for each metabolizer grouping at discharge among patients with WFNS grades of 4-5. mRS indicates modified Rankin scale; WFNS, World Federation of Neurological Surgeons.

## Discussion

There are major clinical implications using a one-size fits all 60mg nimodipine dosage for all patients with aSAH regardless of body weight. A fixed 60 mg every 4 hours does not consider a patient’s individualized body weight, nor initial severity of illness (17), which can lead clinical dose-modification and reduction. A reduction in standard 60mg dosage of nimodipine is critically important since it is the only FDA-approved neuroprotectant shown in several randomized controlled trials to improve clinical outcomes. Understanding the drug’s pharmacogenomic metabolism may further elucidate a future precision medicine approach compared to a one-size fits all approach.

As Figure 2 illustrates, an individualized dosing of nimodipine for patients with aSAH might be considered, since our patients had different responses to the initial 60-mg dose. Further, among the randomized, controlled nimodipine trials, only Allen et al (8) used an initial weight-based 0.7 mg/kg dose (rounded to the nearest 10-mg capsule), followed by a maintenance dosage of 0.35 mg/kg every 4 hours for up to 21 days. For example, a 70-kg patient would initially receive a 50-mg dose (0.7 mg/kg ×70 =49 mg rounded up), followed by 24.5 mg every 4 hours.

Nimodipine’s package insert states that hypotension was reported in about 5% of patients during these historical clinical trials. This appears inconsistent with more recent observations by Wessell et al (7) and our own population, where more than half had hypotension after the 60 mg dose. Theoretically, there may have been a lower incidence of hypotension in these trials if these patients had severe underlying hypertension at aSAH onset, providing better tolerance to nimodipine’s initial potential relative hypotensive effect. Another possibility is that in today’s neurologic intensive care unit there may be increased detection of relative hypotensive events with the use of arterial line systolic blood pressure monitoring.

The results in Figure 2 suggest that there are different nimodipine dosages used over time after the initial 60 mg dose, including dose reduction to 30 mg, 15 mg, or even briefly stopping the drug. One question raised by this observation relates to the maximal dose of nimodipine that can be tolerated. Allen et al (8) mentioned that based on nimodipine’s affinity to the calcium receptor and on levels detected in the cerebrospinal fluid (CSF), a dose 3 to 6 times higher could be theoretically considered. Petruk et al (9) used a 90-mg dose in their trial, maximal dose higher than the one employed in our cohort.

Our data show that patients with lower WFNS grades at admission had more favorable mRS scores throughout all dosing regimens at discharge (Figure 3A), as expected. This group displayed a nonsignificant intergroup decrease as the dose reduced from 60 mg to 30 mg to 15 mg in the graph. For high WFNS grades (Figure 3B), outcomes were generally less favorable (higher proportions of mRS scores, 4-6) because patients were sicker on admission. Interestingly, there was a trend in which a higher rate of patient mortality was seen as the dose of nimodipine decreased. This warrants further study because although patients in this group were admitted sicker and may have been less tolerant to drugs overall, the highest proportion of deaths in the low-grade WFNS group was also seen with the 15-mg dosing regimen. While our results do not indicate a highest effective dose for nimodipine, they suggest a minimally effective dose closely below 30 mg.

Figure 4 demonstrates percentage differences in mRS between the metabolizer groups. While our data yielded no significant results, there was a trend of more favorable mRS scores in the extensive metabolizers group compared to the poor metabolizers (Figure 4A). Further, we observed that the percentage of deaths within the extensive, intermediate, and average metabolizer groups were similar, but almost doubled in the poor metabolizer group (Figure 4B). As shown by Table 3, poor metabolizers have a higher mean age than for the rest of the groups, while extensive metabolizers have the lowest mean age. This could be one of the factors that affects differences in outcome. Determine causation solely between genotype and outcomes is complex. Our data are underpowered to detect significant differences in outcomes; only a properly-powered, prospective, randomized clinical trial could assess this issue.

One of the limitations of our study is the existence of numerous factors that interplay and may alter a patient’s metabolism. Phenoconversion is a concept that embodies the discrepancies between genotype-predicted drug metabolism and the patient’s true capacity to metabolize drugs (18). Klomp et al (18) reports that phenoconversion into lower metabolizer genotypes by, for example, CYP450 inhibitors, inflammation, or age, is possible. As mentioned, in Table 3, we can see that the poor metabolizers correspond to a higher mean age, as opposed to the extensive metabolizers that correspond to a lower mean age, a difference that is statistically significant. Hence, consideration of these variations, both extrinsic and intrinsic, should be incorporated into models that aim to develop precision medicine approaches.

We acknowledge another limitation of this study is its retrospective design. However, our study is the largest pharmacogenomic study of nimodipine in aSAH to date. Further, our data provides real-world observations of dose reduction of nimodipine seen in clinical practice that likely occur at other comprehensive stroke centers who manage complex aSAH patients. Our rate of adverse drug reaction dose reduction was 57% (86/150) which is similar to that reported by Wessell et al (7), who reported as high as 78% dose intolerance due to hypotension from the standard nimodipine dose of 60 mg every 4 hours. The electronic health record did capture dose reduction in the medication administration record effectively in the neuro-intensive care unit.

Dose reduction from the standard 60 mg nimodipine dose is an important clinical finding since it was associated with worse neurological outcomes, particularly when less than 30 mg. Sandow et al (6) also observed that nimodipine-induced relative hypotension can reduce cerebral perfusion pressure which is concerning in this critically ill population because it can lead to a decrease in cerebral blood flow. This may be a potential explanation for worse clinical outcomes seen with lower nimodipine dose in our data and Sandow et al (6) point out this is seen in poor-grade aSAH patients. Physiologically, hypotension is a complex, multifactorial response variable and one that warrants a prospective multimodal approach to capture multiple covariates in conjunction with pharmacogenomics.

Patients with poor-grade aSAH have higher Hunt Hess (HH) (19) and WFNS grades (20) at onset. These higher HH and WFNS grades indicate worse overall initial clinical severity. Therefore, patients with poor-grade aSAH are sicker physiologically and more prone to hemodynamic instability and may be intolerant to the standard dose of 60mg. Poor-grade aSAH patients may be more susceptible to hypotension, have systemic inflammatory response syndrome, early sepsis, metabolic acidosis, and/or reduced left ventricular ejection fraction dysfunction such as takotsubo cardiomyopathy (21). A specific scale such as the Physiological Derangement Scale (17) may be useful to assess the patient’s physiological state at admission.

An important observation in a small subset of patients whose dose was reduced demonstrated non-significant neurologic improvement when the nimodipine dose was later increased back to the standard dose compared to patients still receiving reduced doses at discharge. This again raises the question about there being a potential minimally effective dose for nimodipine neuroprotection of 30 mg every 4 hours, like the maintenance dose of 37.5mg/kg every 4 hours proposed by Allen et al (8). Furthermore, perhaps a higher tolerable dose is possible with sufficient pharmacogenomic metabolism of nimodipine as seen in clopidogrel genotype-driven cardiac intervention trials (22).

There are complex drug-drug interactions with the CYP system (23) or changes in CYP system in older patients (24, 25) taking nimodipine. For example, dexamethasone, which is sometimes given to neurological and neurosurgical patients with craniotomy and clipping of a ruptured brain aneurysm, can cause a drug-drug interaction with nimodipine and would theoretically raise serum levels of nimodipine, whereas phenytoin can reduce nimodipine levels. While we did not measure concomitant blood and CSF levels of the nimodipine with pharmacogenomics types in this study to account for drug-drug interactions on nimodipine levels, we separately conducted a small Phase I prospective pilot of comprehensive pharmacogenomic genotype-phenotype correlations that included samples of blood and CSF nimodipine levels but was distinct from this larger cohort.

While our study population included 34 Black patients (22.7%), studies with greater racial, ethnic, and pharmacogenomic genetic diversity will be warranted in the future. Female sex is well represented in this study because historically aSAH is a 1.5:1 or 2:1 female-to-male predominant disease (26).

Gut absorption of nimodipine is another important consideration in patients with poor grade aSAH. In a study by Abboud et al (27) comparing nimodipine levels with enteral versus intravenous (IV) administration, levels were lower with enteral administration. While there are numerous factors to consider, in the intensive care setting, patients may develop gastrointestinal ileus, causing accumulation of the drug in the stomach or proximal intestine which may in turn cause impaired gut and hepatic CYP metabolism of the drug. These gut changes may lead to different drug bioavailability or relative intestinal malabsorption issues within the gut (28). Past medical history of liver disease was limited to only 4 patients, and liver function tests were not done for most patients (only 18 [12%] had elevated test results).

Abboud et al (27) noted that levels of nimodipine were higher in patients with higher WFNS grades receiving IV administration compared to enteral. This is important because IV administration bypasses gut absorption and first-pass hepatic CYP3A4 and CYP3A5 metabolism (29). While IV nimodipine is allowed in Europe, it is illegal in the US due to an FDA black box warning (30) issued after reports of cardiovascular instability and death (5) and because IV administration involves injecting liquid extracted from the enteral capsules. This is no longer the case for the IV formulation available in Europe.

The FDA reported through its Adverse Event Reporting System and other sources that 25 of 31 cases of nimodipine errors between 1989 and 2009 were due to IV injection of liquid from nimodipine capsules. Four patients given this IV administration died and 5 experienced serious or near-fatal harm. Therefore, enteral administration of nimodipine (via nasogastric tube or equivalent and flushed with 30 mL of normal saline [0.9%] for patients unable to take by mouth) remains the only form available in the US. This coupled with the FDA-recommended 60-mg initial and maintenance dose enterally every 4 hours for 21 days make it challenging to treat patients who are intolerant —or develop relative hypotension while receiving—the drug, regardless of the underlying cause.

The genotypes of patients who experienced the severe adverse drug events reported by the FDA after IV administration of enteral nimodipine remain unknown. However, the answer to why IV nimodipine can be given in Europe but not the US likely remains around the specific formulation and method of administration and may be beyond basic pharmacogenomics status. For most drugs in the US in critically ill patients, IV administration is a standard backup option when enteral intolerance and gut absorption are major concerns and enteral nutrition must be held when surgery is planned. Additionally, worse outcomes in patients with poor-grade aSAH may be due not only to poor gut absorption of the drug, but also altered brain-gut axis, altered gut microbiome, and factors which are poorly understood that indirectly alter the underlying drug pharmacodynamic model (31).

## Conclusion

This is the largest-known retrospective analysis of the pharmacogenomics of nimodipine on clinical outcomes for patients with aSAH. Overall, patients who tolerated the standard dose of 60 mg every 4 hours had increased percentages of better outcomes compared to those on lower doses and those who had their dose reduced then increased back to 60 mg. A current pharmacogenomics investigation is underway to better understand how the interplay between blood and CSF drug levels, dosing regimen and genotype might predict risk of hypotension or other adverse events. A larger prospective pharmacogenomics exploration of nimodipine’s dose-response relationship is needed and whether a precision medicine dosing approach could optimize neuroprotection on aSAH patient outcomes.

## Data Availability

All data produced in the present work are contained in the manuscript

## Reprints

W. David Freeman, MD, Department of Neurologic Surgery, Mayo Clinic, 4500 San Pablo Rd, Jacksonville, FL 32224 (Phone 904-956-3331 Fax 904-953-0760)

Presented in part at Mayo Clinic’s 14^th^ Annual Stroke and Cerebrovascular Disease Review Continuous Professional Development course, Amelia Island, FL, Sept 23, 2022.

## Acknowledgment

The Scientific Publications staff at Mayo Clinic provided copyediting, proofreading, administrative, and clerical support.

